# Suicide Death Predictive Models using Electronic Health Record Data

**DOI:** 10.1101/2024.09.26.24314402

**Authors:** Shweta Srikanth, Lina M. Montoya, Matthew M. Turnure, Brian W. Pence, Naoko Fulcher, Bradley N. Gaynes, David B. Goldston, Timothy S. Carey, Shabbar I. Ranapurwala

## Abstract

In the realm of medical research, particularly in the study of suicide risk assessment, the integration of machine learning techniques with traditional statistics methods has become increasingly prevalent. This paper used data from the UNC EHR system from 2006 to 2020 to build models to predict suicide-related death. The dataset, with 1021 cases and 10185 controls consisted of demographic variables and short-term information, on the subject’s prior diagnosis and healthcare utilization. We examined the efficacy of the super learner ensemble method in predicting suicide-related death leveraging its capability to combine multiple predictive algorithms without the necessity of pre-selecting a single model. The study compared the performance of the super learner against five base models, demonstrating its superiority in terms of cross-validated negative log-likelihood scores. The super learner improved upon the best algorithm by 60% and the worst algorithm by 97.5%. We also compared the cross-validated AUC’s of the models optimized to have the best AUC to highlight the importance of the choice of risk function. The results highlight the potential of the super learner in complex predictive tasks in medical research, although considerations of computational expense and model complexity must be carefully managed.

## Introduction

Suicide is a serious public health problem that can have lasting harmful effects on individuals, families, and communities. In 2020, suicide was the second leading cause of death among individuals between the ages of 10-14 and 25-34, the third leading cause of death among individuals between the ages of 15-24, and the fifth leading cause of death among individuals between the ages of 35 and 44. The total age-adjusted suicide rate in the United States increased 35.2% from 10.4 per 100,000 in 2000 to 14.2 per 100,000 in 2018, before declining to 13.9 per 100,000 in 2019 and to 13.5 per 100,000 in 2020 and once again increasing to 14.0 per 100,000 in 2021.^1^

The suicide trends in North Carolina (NC) are similar to those of the nation overall, with increases from about 13 suicide deaths to 16 suicide deaths per 100,000 from 2012 to 2018. In NC, suicide is the third leading cause of death among those aged 5–44 years. Suicide mortality rates vary by gender, race, and age.^2,3^

Risk factors associated with suicide are multifaceted, involving a mix of genetic, biological, clinical, physiological, environmental, and economic factors, among others. The main risk factors for suicide mortality identified in the meta-analyses conducted by Brent et al.^4^ include a previous suicide attempt, suicidal ideation, psychiatric disorders (such as mood and psychotic disorders), physical illnesses (such as cancer and epilepsy), socio-demographic factors (including unemployment and low education), contact with the criminal justice system, state care in childhood, access to firearms, and parental death by suicide. These risk factors were found to be associated with an elevated risk of suicide mortality, with varying degrees of association. The study also highlighted the multi-factorial nature of suicide mortality and emphasized the need for prevention strategies that account for these identified risk factors and their relative strengths.

Considerable research has investigated the relationship between individual predictors and the risk of suicide mortality. Mental health and substance use disorders are recognized as primary predictors of suicide and suicidal tendencies. For example, studies on suicide deaths show that individuals with borderline personality disorder have a standardized mortality ratio (SMR) of 45.1 (95% Confidence Interval [CI] 29.0 to 61.3), while those with opioid use disorder have an SMR of 13.5 (95% CI 10.5 to 17.2).^5–7^ A review by Stack et al.^8^ provides a comprehensive analysis of the environmental factors associated with suicide, highlighting the impact of political, social, cultural, and economic influences. It emphasizes the protective role of social institutions such as marriage, parenting, and religiousness, as well as the influence of economic factors such as unemployment and low income on suicide risk. Furthermore, research involving patients from US health maintenance organizations, notably Kaiser Permanente,^9^ highlighted traumatic brain injury as a significant physical health risk factor for suicide, with an adjusted odds ratio (OR) of 8.80 (95% CI 7.37 to 10.50). While many studies have identified different risk factors linked to self-harm and suicide, focusing on these factors alone does not offer much guidance for clinicians on how to identify those most at risk for suicide. Importantly, it was found that 54% of individuals who died by suicide had not been diagnosed with any mental health condition, indicating that even with notable SMRs linked to mental health issues, these alone do not account for the majority of suicide cases. This suggests a gap in the identification of mental health disorders, highlighting the complex nature of suicide prevention efforts^10–13^

Several existing studies indicate that healthcare utilization tends to rise just before an individual dies by suicide, suggesting that the healthcare system could play a crucial role in suicide prevention efforts.^14–18^ In a national cohort study in Sweden Crump et al.^19^ found that 30.4% and 52.3% of individuals with drug use disorders (DUD) who died by suicide had visited healthcare facilities within 2 weeks and 3 months before their death, respectively, in contrast to 5.9% and 24.3% among controls. The research underscores the critical need for recognizing and addressing suicidal tendencies in DUD patients, particularly in primary and specialty outpatient settings, where many of these patients had their last clinical visits shortly before their suicide. The study by Qin et al.^20^ identifies critical suicide risk peaks immediately after psychiatric admission and discharge, noting higher risks for those with shorter hospital stays and significant impacts of affective disorders on suicide rates. The study by Ho et al.^21^ reveals a significantly elevated suicide risk among psychiatric patients in Hong Kong in the year following hospital discharge, particularly within the first 28 days, with the highest risks observed in young adults and females, regardless of specific diagnoses.

However, the populations studied were subject to healthcare systems vastly different from that of the United States. Hence large-scale, longitudinal studies integrating multiple databases and perspectives are essential for identifying predictors of suicide, building prediction models and informing targeted interventions.

There have been several existing studies that have built models for predicting suicide-related outcomes. Building an effective prediction model for suicide-related outcomes requires a nuanced understanding of both epidemiological and clinical aspects of suicide prevention.^22^ In parametric regression, the bias-trade variance trade-off is pronounced. A logistic regression may overfit the data if there are more parameters than observations. But if a model is too simple, the results may not accurately capture the predictor-outcome relationship. In this context, flexible machine learning procedures offer a promising alternative, allowing researchers to uncover intricate relationships among suicide risk factors, with safeguards against an overfit. With a growing abundance of data, there is an increasing interest in utilizing machine learning for developing suicide prediction models across various populations.^23–29^ But the choice of the most appropriate model for the data remains a challenge.

Ensemble machine learning techniques provide researchers with the flexibility to explore multiple algorithms simultaneously. These methods, combine predictions from various base learners to create a more robust and accurate model. By incorporating diverse algorithms into the ensemble, researchers can leverage the strengths of each individual method while mitigating their weaknesses. This approach allows for a comprehensive exploration of different modeling approaches, including both traditional statistical methods and more complex machine learning algorithms. By employing ensemble learning techniques, these models offer valuable insights into the intricate interplay of suicide risk factors. Consequently, while statistical advancements show promise for enhancing suicide risk assessment, a nuanced understanding of model capabilities and limitations is essential for informed clinical decision-making and the development of effective suicide prevention strategies.

The paper follows this structure: Section 2 provides details on the dataset and methods, Section 3 presents the results, and Section 4 discusses the findings.

## Methods

### 0.1 Participants

The current study^30^ links mortality information from 2006-2020 from two comprehensive statewide systems, the NC death certificate data and The North Carolina Violent Death Reporting System (NCVDRS), to four different target subpopulations:

1. electronic health records of a large healthcare system serving residents of all 100 counties of the state
2. claims records of a large single private health insurance carrier in the state
3. claims records of the state Medicaid program
4. justice individuals released from the NC prisons following incarceration.

For the scope of this paper, our focus will be on the data associated with UNC EHR exclusively. Death certificates from 2006 to 2022, publicly accessible records managed by the NC Department of Health and Human Services (DHHS), are utilized. NCVDRS, a state-based surveillance system, collates information from various sources including law enforcement, coroners, medical examiners, vital statistics, and crime laboratories to compile detailed records of violent deaths within each state. The study incorporates NCVDRS data spanning from 2006 to 2022. The UNC electronic health records (UNC EHR) encompass data from the UNC Health System, which delivers medical care to over 1 million patients annually, constituting approximately 10% of the North Carolina population, across all 100 counties.

The study employed a nested case-control design, where the total population is those within the UNC Healthcare system. The cases and controls are sampled from within a risk set. A person enters the risk set when they accrue 3 months of having been in care at the UNC Healthcare System, they leave the risk set if they go *≥* 3 months without any encounter in the UNC Healthcare System. Case events are all suicide deaths as defined by ICD codes, from within the risk set. Control Events are randomly selected per Case Event. Controls are matched to Case Events based on inclusion in the respective monthly Risk Set, as well as factors such as sex, and age group. Random selection is without replacement per Case Event and with replacement across Control Events.

Selecting controls based on incidence density-matched controls addresses significant selection bias concerns often encountered with convenience or high-risk control groups commonly found in suicide literature.^31^ These cases and controls will then be utilized to formulate a series of predictive algorithms aimed at identifying individuals at risk of short-term suicide death.

### 0.2 Covariates and Outcome

The outcome of suicide death is identified using ICD 10 codes (U03, X60–X70, X71–X83 and Y87.0) via linkage of NC death records and NCVDRS with UNC EHR data. The primary covariates of interest include demographics like age, sex, and race, healthcare utilization and prior diagnosis. We are specifically interested in short-term information, which we define as data captured within one month leading up to the event.

Diagnosis variables are categorized into four groups: acute, sub-acute, chronic, and permanent diseases. The acute conditions such as acute pain contain one-month prior information. For sub-acute conditions, we add 6 additional months of history, for chronic conditions we add 12 additional months of history and for permanent health conditions, we use all avail-able history. Each diagnosis has 3 variables with information about whether the person had any encounters, number of encounters and number of unique encounters in the healthcare system related to that diagnosis in the defined time period. Healthcare utilization variables include flag variables to indicate if a person has had any inpatient, outpatient or emergency room visits, number of inpatient, outpatient, or emergency room visits, and total number of days spent in the hospital as an inpatient in the past 1 month. This gives us a total of 48 covariates.

### 0.3 Model for Predicting Suicide Deaths

The primary aim is to predict the risk score of suicide death. Our statistical task is to estimate the probability of suicide death, given the covariates measured in the study. The method used in this paper is a super learner. Superleaner is an ensembling machine learning approach that combines multiple algorithms into a single algorithm and returns a prediction function with the lowest cross-validated risk of choice such as MSE or expected Negative Log Likelihood.^32^ This approach hence frees investigators from the need to commit to a single regression technique from the beginning; instead, it allows them to simultaneously employ multiple methods. By leveraging several relevant algorithms, researchers are able to determine the most effective function tailored to their specific data set. Thus with Super Learning, several algorithms can“work together” to create a combination of algorithms that can outperform any single candidate algorithm in the library (including simple, parametric regressions or more data-adaptive algorithms such as random forests or neural networks). Figure 1 describes the super learner algorithm.

**Figure 1:**
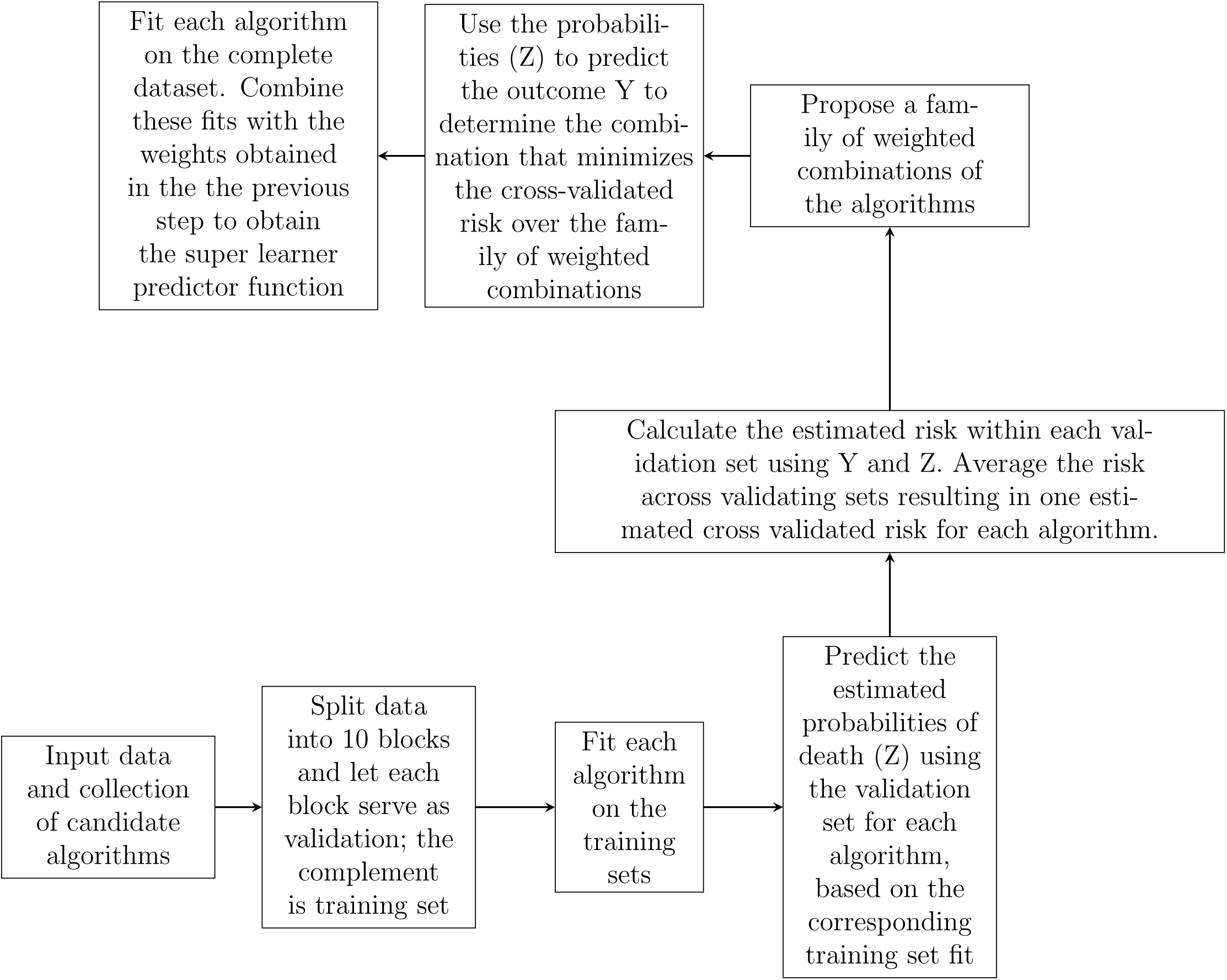
Superlearner algorithm flowchart.

We define the full data structure as *X* = (*W, Y*) *∼ P_X,_*_0_ with j covariates *W*_1_*, …, W_j_* and binary outcome *Y*, indicating suicide death. The full data structure refers to all subjects in the risk set. We introduce a random variable *O* to represent the observed data, *O* = (*Y,* Δ, Δ*X*) = (*Y,* Δ, Δ(*W, Y*)) *∼ P*_0_ where Δ is an indicator of inclusion in nested case-control sample from the risk set. The target statistical parameter of interest as a function of the of the full data distribution of *X* is given by *Q̅* = *E_X,_*_0_(*Y |W*) and the full-data statistical model denoted by *M^F^* (full data) is non-parametric. *E*_0_*L*(*O*, *Q̅*) is the expected loss, which evaluates the candidate *Q̅*.

We choose negative log likelihood as the loss function, represented by *L^F^* (*X*, *Q̅*). If our sample had consisted of n i.i.d. observations *X_i_*, we would estimate *Q̅* = *E_X,_*_0_(*Y |W*) simply using the loss function *L^F^* (*X*, *Q̅*). However, given that we observe O, we estimate *Q̅* with super learning and weights 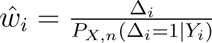 for observation *i* = 1*, …n*, which now gives us an inverse probability weighted loss function with input *X* :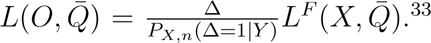 Thus our target parameter of interest is defined as *Q̅* = arg min*_Q̅_ E*_0_*L*(*O*, *Q̅*)) where *Q̅* is a possible function that is compatible with our current prediction problem. *Q̅* is minimized at the optimal choice of *Q̅*_0_.

The super learner algorithm is weighted due to the nested case control study design. The probability of being sampled in the nested case-control study, given that a person experienced suicide death= 1 because all eligible cases in the risk set are sampled. The probability of being sampled in the nested case-control study, given that a person did not experience suicide death is calculated as the Total number sampled as controls/ Total number at risk of being sampled as controls, which was equal to 0.000131. We implement super learning in the R programming language^34^ with the SuperLearner package^35^ to estimate *Q̅*_0_.

In the construction of our predictive model, we used a diverse library of candidate learners to ensure robustness and accuracy. We only use candidate learners that allow for the incorporation of weights. This library included: the mean algorithm, generalized linear models, elastic net penalized linear regression models, generalized additive models, and recursive partitioning and regression trees. Each learner was chosen for its unique statistical properties and strengths in handling different types of data structures and relationships, facilitating a comprehensive approach to prediction. We restricted to these 5 models due to computational limitations.

## Results

The summary of the dataset can be found in Tables 1 and 2. Of the 11,206 observations, 1021 are cases with a suicide death and 10,185 are controls without an event. 72.4% of the observations are male, 68.7% are white, and the mean age is 51.58 with a standard deviation of 17.91. The tables also contains information about the person’s prior diagnosis and healthcare utilization information. There are a total of 48 variables, and we present a select few variables in the tables. The most prevalent diagnoses are cardiovascular diseases, mental health and chronic pain. Around 15% of the subjects had an outpatient visit, 0.3% had an emergency room visit and 1.1% had an inpatient visit in the month before the event.

**Table 1:** Descriptive Statistics of Categorical Variables.

| **N=11206** | **n(%)** |
| --- | --- |
| Case | 1021 (9.11) |
| Controls | 10185 (90.89) |
| **Sex** |  |
| Male | 8108 (72.40) |
| Female | 3098 (27.60) |
| **Race** |  |
| White | 7704 (68.70) |
| Black | 2019 (18.00) |

Table 1: Descriptive Statistics of Categorical Variables
| **N=11206** | **n(%)** |
| --- | --- |
| Asian | 188 (1.70) |
| Native American | 93 (0.80) |
| Other | 1071 (9.60) |
| Acute pain in the last 1 month | 202 (1.80) |
| Surgery in the last 7 month | 346 (3.10) |
| Mental health encounters in the last 13 months | 1422 (12.7) |
| Chronic pain encounters in the last 13 months | 3534 (31.5) |
| Cardiovascular encounters in the last 13 months | 4124 (36.8) |
| Inpatient encounter in the last 1 month | 122(1.10) |
| ER visit encounter in the last 1 month | 33 (0.30) |
| Outpatient encounter in the last 1 month | 1701 (15.20) |

**Table 2:** Descriptive Statistics of Continuous Variables.

| **N=11206** | **mean(sd)** |
| --- | --- |
| Age | 51.58 (17.91) |
| Number of acute pain related encounters in the last 1 month | 0.07 (0.64) |
| Number of unique acute pain related encounters in the last 1 month | 0.04 (0.26) |
| Number of surgery related encounters in the last 7 months | 0.11 (2.58) |
| Number of unique surgery related encounters in the last 7 months | 0.04 (0.49) |
| Number of mental health related encounters in the last 13 months | 0.37 (1.78) |
| Number of unique mental health related encounters in the last 13 months | 0.21 (0.69) |
| Number of Chronic pain related encounters in the last 13 months | 1.07 (3.28) |
| Number of unique Chronic pain related encounters in the last 13 months | 0.97 (2.32) |
| Number of Cardiovascular related encounters in the last 13 months | 1.28 (3.22) |

Table 2: Descriptive Statistics of Continuous Variables
| **N=11206** | **mean(sd)** |
| --- | --- |
| Number of unique Cardiovascular related encounters in the last 13 months | 0.90 (1.75) |
| Number of inpatient encounter in the last 1 month | 0.01 (0.11) |
| Number of total inpatient hospital stay in the last 1month | 0.06 (0.74) |
| Number of ER visit encounter in the last 1 month | 0 (0.10) |
| Number of outpatient encounter in the last 1 month | 0.31 (1.13) |

Before running the model we removed continuous variables with 0 variablilty and remove categorical variables with less than 1% representation in the dataset.

Observation weights within the super learner were assigned based on the inverse proba-bility of missingness, 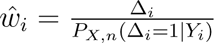. Thus cases were given observation weights equal to 1 and controls were given observation weights of 1/0.000131 = 7633.588. This number was calculated based on the monthly risk set information and the number of cases and controls in the dataset.

Table 3 presents candidate learners and libraries used in the model. The negative log-likelihood for each algorithm, along with the calculated weight associated with each algorithm in the ensemble SL model is presented in Table 4. The highest weight of 0.753 is associated with recursive partitioning and regression trees, followed by a weight of 0.119 for generalized additive model.

**Table 3:** Candidate Learners.

| **Model** | **Library** |
| --- | --- |
| Mean | SL.mean |
| Main Terms Logistic Regression | SL.glm |
| Penalized regression using elastic net. | SL.glmnet |
| Generalized Additive Model | SL.gam |
| Recursive partitioning and regression trees | SL.rpart |

**Table 4:**
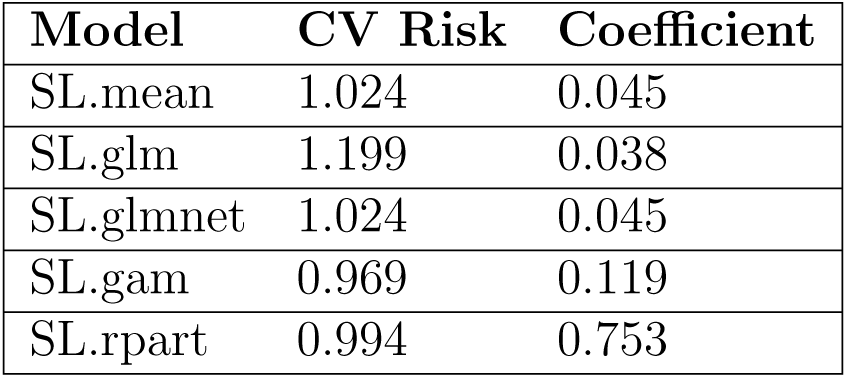
CV NLL and Coefficient of each candidate.

| **Model** | **CV Risk** | **Coefficient** |
| --- | --- | --- |
| SL.mean | 1.024 | 0.045 |
| SL.glm | 1.199 | 0.038 |
| SL.glmnet | 1.024 | 0.045 |
| SL.gam | 0.969 | 0.119 |
| SL.rpart | 0.994 | 0.753 |

Figure 2 represents the minimum, average, and maximum negative log-likelihood of each model.

**Figure 2:**
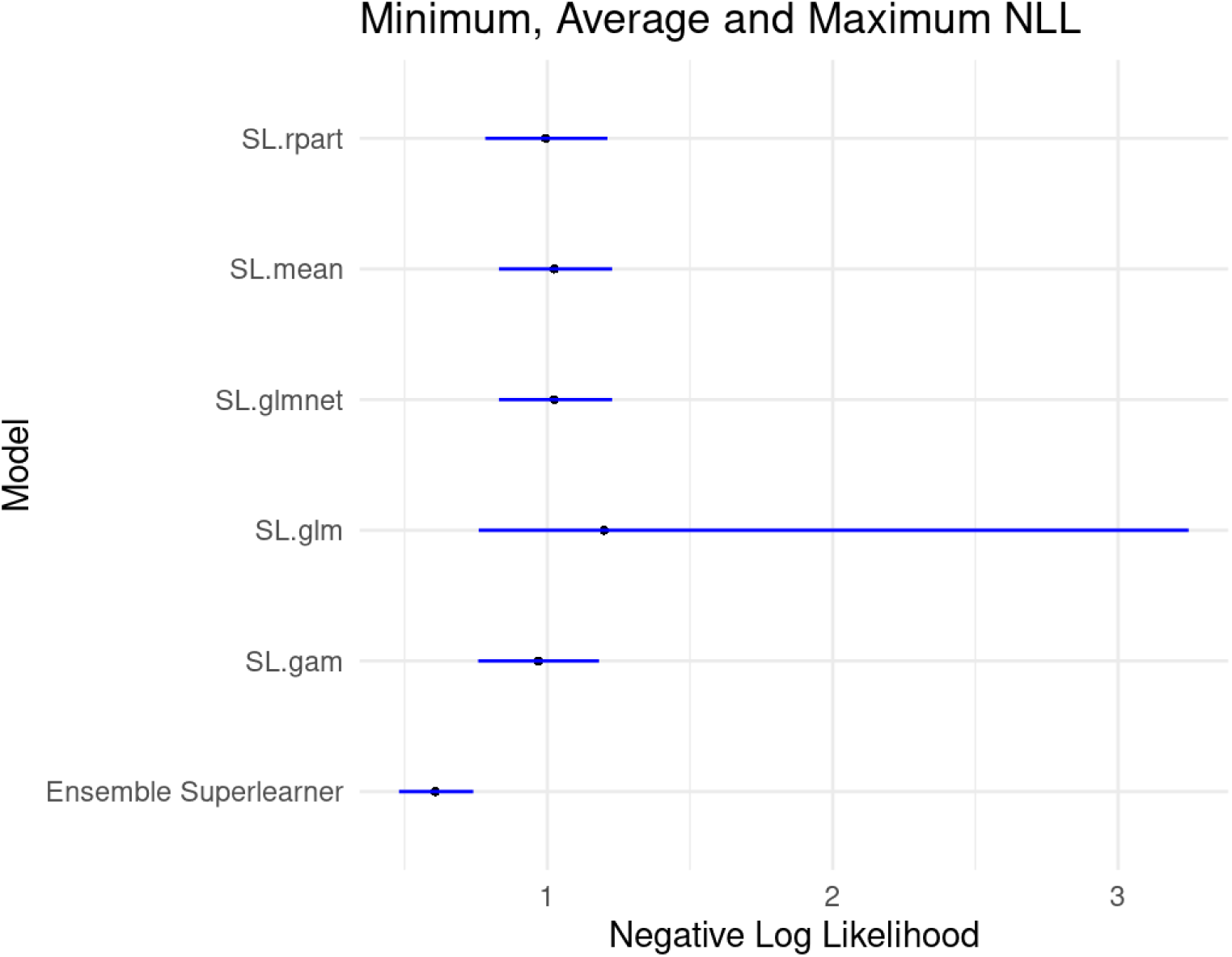
10 fold CV Risk Estimate of models.

Table 5 presents the Relative Efficiency with respect to negative log-likelihood for each algorithm. Relative efficiency for each of the k algorithms was defined as RE = CV NNL(k)/CV NNL(super learner). Superlearner improved upon the best algorithm generalized additive models by around 60%. The super learner improved upon the worst algorithm, main terms logistic regression by 97.5%. We also fit another model, with the same 5 base learners, but this time used AUC as the risk function so that they are optimized to have the best AUC. In Table 6 we present the cross-validated AUC of the different base learners and the ensemble superlearner. In this case, ensemble SL has the highest AUC of 0.74, with the other base learners having AUC’s ranging from 0.5 to 0.72.

**Table 5:** Relative Efficiency With Respect to Cross-Validated NNL.

| **Model** | **CV NLL** | **Relative Efficiency** |
| --- | --- | --- |
| Ensemble Superlearner | 0.607 | - |
| SL.mean | 1.024 | 1.687 |
| SL.glm | 1.199 | 1.975 |
| SL.glmnet | 1.024 | 1.687 |
| SL.gam | 0.969 | 1.596 |
| SL.rpart | 0.994 | 1.638 |

**Table 6:** CV AUC for model with AUC as Risk fucntion.

| **Model** | **CV AUC of Model with AUC as risk** |
| --- | --- |
| Ensemble Superlearner | 0.74 |
| SL.mean | 0.5 |
| SL.glm | 0.71 |
| SL.glmnet | 0.5 |
| SL.gam | 0.72 |
| SL.rpart | 0.51 |

## Discussion

Research in medicine and epidemiology has focused significantly on developing functions for predicting risk scores, aiming to generate risk assessments for various diseases. In the context of suicide, several studies have developed models for predicting suicide-related outcomes. Apart from traditional statistics methods, there has been a growing interest in integrating machine learning techniques, owing to the increasing abundance of data. However, determining the superior algorithm for a given dataset remains a challenge prior to analysis. The primary advantage of using an ensemble algorithm such as the super learner is that it eliminates the need to choose a single algorithm a priori. This method allows for the simultaneous application of multiple candidate learners to identify an enhanced, optimal estimator. It also facilitates the selection of the most appropriate loss function for optimizing the algorithm, and employs cross-validation to prevent overfitting. In this study, we chose the Negative Log Likelihood because our main interest lies in accurately predicting probabilities rather than merely distinguishing between events, for which the AUC (Area Under the Curve) would be a more suitable choice.

The ensemble super learner had the best performance compared to all 5 models. The model significantly improved upon all the base learners with respect to CV Expected Non-Negative Log Likelihood. Even in the case when the improvement from the base learners is small, superlearner is still advantageous because it can provide researchers with the ability to choose the best model, without needing to commit to any single model a prior. The choice of the risk function is critical and must be based on the scientific question of interest. In our case, the focus is on risk score prediction, which relies on accurately predicting the probabilities rather than merely distinguishing between the events. In this case, Negative Log Likelihood is a better metric than AUC.

The major drawback with implementing super learner is the computational time and intensity when compared to running a single model. This becomes further more a constraint with increasing number of base learners. The success of super learner is contingent upon the base learners that are provided. Ideally, one would use all relevant algorithms as base learners to find the most optimal model. However, due to above mentioned constraints, there is often a limitation on the number of base learners one specifies. Hence the decision regarding which algorithms to use must be systematic and well informed.

For future work, focusing on assessing the importance of variables within the super learner framework could be valuable. Understanding which features contribute most significantly to the predictive performance of the model would prove to be very valuable insights to clinicians.

Prior research has consistently indicated that suicide attempts and suicide ideations are robust predictors of suicide-related death^36^ Furthermore, certain medications have been associated with an increased risk of suicide attempts and fatalities.^37^ To enhance predictive models for suicide risk, it is crucial to incorporate these elements into the analytical framework. By integrating variables such as history of suicide attempts, presence of suicide ideation, and use of high-risk medications, the model can offer a more comprehensive approach to assessing suicide risk.

Relying solely on EHR data also introduces several limitations. The accuracy and reliability of EHR data can vary significantly depending on the quality and meticulousness of data entry practices.^38^ Additionally, EHRs may not capture non-clinical factors that significantly contribute to suicide risk, such as socioeconomic status or life stressors, which are often not documented systematically. Therefore, while the integration of EHR data enhances predictive modeling for suicide risk, it is essential to acknowledge and address these limitations to develop more comprehensive and effective risk assessment tools in the field of suicide prevention.

## Conclusions

In this study, we applied the super learner ensemble algorithm to predict suicide-related outcomes. Our analysis demonstrates the effectiveness of ensemble learning in integrating diverse algorithms, notably recursive partitioning and regression trees, to enhance predictive accuracy. Despite computational challenges, the super learner offers a systematic approach to model selection and optimization, resulting in significant improvements over individual base learners.

Looking ahead, the super learner algorithm presents a promising avenue for enhancing predictive modeling in healthcare research. Future efforts should concentrate on refining this algorithm to overcome computational limitations and unlock its full potential. It will also be important to explore the development of clinically relevant tools, such as identifying which variables are most predictive of outcomes. Overall, our study highlights the significant value of ensemble techniques, especially the super learner, in improving risk assessments and aiding clinical decision-making processes.

## Data Availability

The data are not available upon request to the authors.

## Notes

### Competing Interest Statement

The authors have declared no competing interest.

### Funding Statement

This study was funded by a grant from the National Institute of Mental Health R01MH124752 (Pence and Ranapurwala, MPI)

### Author Declarations

IRB of University of North Carolina gave ethical approval for this work.

